# Development of medical device software for the screening and assessment of depression severity using data collected from a wristband-type wearable device: SWIFT study protocol

**DOI:** 10.1101/2022.03.09.22271883

**Authors:** Taishiro Kishimoto, Shotaro Kinoshita, Toshiaki Kikuchi, Shogyoku Bun, Momoko Kitazawa, Toshiro Horigome, Yuki Tazawa, Akihiro Takamiya, Jinichi Hirano, Masaru Mimura, Kuo-ching Liang, Norihiro Koga, Yasushi Ochiai, Hiromi Ito, Yumiko Miyamae, Yuiko Tsujimoto, Kei Sakuma, Hisashi Kida, Gentaro Miura, Yuko Kawade, Akiko Goto, Fumihiro Yoshino, the SWIFT collaborators

## Abstract

**Introduction:** Few biomarkers can be clinically used to diagnose and assess the severity of depression. However, a decrease in activity and sleep efficiency can be observed in depressed patients, and recent technological developments have made it possible to measure these changes. In addition, physiological changes, such as heart rate variability, can be used to distinguish depressed patients from normal persons; these parameters can be used to improve diagnostic accuracy. The proposed research will explore and construct machine learning models capable of detecting depressive episodes and assessing their severity using data collected from wristband-type wearable devices.

**Methods:** Patients with depressive symptoms and healthy subjects will wear a wristband-type wearable device for 7 days; data on triaxial acceleration, pulse rate, skin temperature, and ultraviolet light will be collected. On the seventh day of wearing, the severity of depressive episodes will be assessed using SCID-5, HAMD, and other scales. Data for up to five 7-day periods of device wearing will be collected from each subject. Using wearable device data associated with clinical symptoms as supervisory data, we will explore and build a machine learning model capable of identifying the presence or absence of depressive episodes and predicting the HAMD scores for an unknown data set.

**Discussion:** Our machine learning model could improve the clinical diagnosis and management of depression through the use of a wearable medical device.

**Trial Registration:** jRCT1031210478, Japan Registry of Clinical Trials (jRCT)

## 1. INTRODUCTION

Currently, few biomarkers can be clinically used to diagnose and assess the severity of mental disorders.^1,2^ For this reason, psychiatrists must diagnosis and/or assess severity based on interviews with patients. However, such evaluation methods, which are influenced by the patient’s perception and the clinician’s experience and senses, may lower the diagnostic agreement rate and make it difficult to determine the effect of treatment.

In recent years, studies using accelerometer-equipped actigraphs have been conducted to quantify the sleep disturbances and decreased activity levels that often occur in patients with mental disorders.^3-5^ However, most of these studies were unable to determine the risk of depression or to assess the severity of depression in individual patients.^6^

On the other hand, the latest wearable devices are capable of previously unavailable measurement modalities, such as heart rate, respiration rate, skin temperature, and location, in addition to actigraphy functions similar to those used for research, and research on depression is now being conducted using these new devices.^7,8^ Our research group previously reported that depressed patients could be identified and the severity of depression could be estimated with a certain degree of accuracy by applying machine learning to data obtained from 86 depressed and healthy people who wore a wristband-type wearable device for 7 days.^9^ Although these results were preliminarily based on a limited number of subjects, we believe that this machine learning approach could reach a clinically useful level once more data has been accumulated. Specifically, in the learning model used in our previous study, sleep, body movement, skin temperature, the correlation between sleep and skin temperature, and heart rate were selected as features.^9^ In addition, since other previous studies have shown that heart rate variability is useful for identifying mood changes and depression,^10,11^ the learning model might be improved by the addition of these new parameters.

In the presently proposed study (SWIFT study: SoftWare development Integrating wearable technologies for Future depression Treatment), we will collect data and build machine learning models for the development of medical device software capable of detecting and quantifying the severity of depressive episodes in subjects with depression and bipolar disorder.

## 2. METHODS

### 2.1. Design Overview

In this study, patients with depressive symptoms (depressive disorder, bipolar disorder and related disorders, anxiety disorders, obsessive-compulsive and related disorders, trauma-and stressor-related disorders, dissociative disorders, somatic symptoms and related disorders) and healthy subjects will be asked to wear a wristband-type wearable device for 7 days. On the last day of the measurement period, a well-trained evaluator will (1) assess whether the subject meets the criteria for a depressive episode as defined by the DSM-5 using the SCID-5, and (2) assess the severity of the depressive episode using the HAMD and other rating scales. These assessments will be used as clinical information and linked with the wearable device data. Data for up to five 7-day periods of device wearing will be collected from each subject. Additionally, structural MRI and resting state fMRI data will be collected for some subjects who consent to these examinations.

Information including triaxial acceleration (which can be converted into information on body movement, sleep, etc.), pulse, skin temperature, and UV exposure will be obtained from the wearable devices. The data will be sent to a secured cloud server via the research collaborator’s smartphone and retrieved by the research group.

### 2.2. Participants

This study will be an open-label observational study. Patients who meet the inclusion criteria will be recruited at the time of their outpatient visit to Keio University Hospital and other medical facilities or during their hospitalization, and healthy individuals without a history of mental illness will be recruited through recruitment advertisements on a website. Patient recruitment will be conducted at the following locations and medical facilities: Tokyo (Keio University, Akasaka Clinic, Oizumi Hospital, Oizumi Mental Clinic, Tsurugaoka Garden Hospital); Kanagawa (Nagatsuta Ikoinomori Clinic); Fukushima (Asaka Hospital); and Yamagata (Sato Hospital). Other medical facilities may be added depending on the recruiting situation.

The inclusion criteria for patients shall be as follows: 1) a clinical diagnosis of depressive disorder, bipolar or related disorder, anxiety disorder, obsessive-compulsive or related disorder, trauma-and stressor-related disorder, dissociative disorder, or somatic symptom or related disorder according to the DSM-5 criteria, the presence of depressive symptoms, and an outpatient or inpatient status at Keio University Hospital or another medical facility participating in the study; 2) an age of 18 years or older at the time of consent; 3) confirmation from a psychiatrist that the subject is capable of providing written consent or that a guardian can provide consent if the psychiatrist determines that obtaining consent from the patient will be difficult; and 4) ownership and ability to use a smart phone (iOS 11. 0 or later or Android version 5.0 or later). The inclusion criteria for healthy subjects shall be as follows: 1) no history of psychiatric illness and cooperation with the study on a volunteer basis (after obtaining consent, the M.I.N.I. will be used to confirm that the subject does not have a psychiatric illness); 2) an age of 18 years or older at the time of consent; and 3) ownership and ability to use a smartphone (iOS 11.0 or later or Android version 5.0 or later).

The exclusion criteria for patients shall be as follows: 1) an inability to complete measurements that could affect their medical conditions, as determined by a psychiatrist; 2) the presence of comorbidities that could affect the measurement results, such as paralysis of the upper limbs; and 3) ineligibility for other reasons determined by the principal investigator or sub-investigator. The exclusion criteria for healthy subjects shall be as follows: 1) the presence of comorbidities that could affect the measurement results, such as paralysis of the upper limbs; and 2) ineligibility for other reasons determined by the principal investigator or sub-investigator. Researchers will obtain written informed consent from all the participants. The participants will be able to leave the study at any time.

### 2.3. Assessment Schedule

The assessment schedule is presented in Table 1. Subjects will be asked to wear a wristband-type wearable device. On the seventh day of wearing the device, (1) the SCID-5 will be used to assess whether the subject meets the criteria for a depressive episode as defined by the DSM-5, and (2) the HAMD will be used to assess the severity of the depressive episode. These assessments will be linked to the wearable device data. A maximum of five 7-day periods of device wearing will be allowed per subject.

**Table 1.**
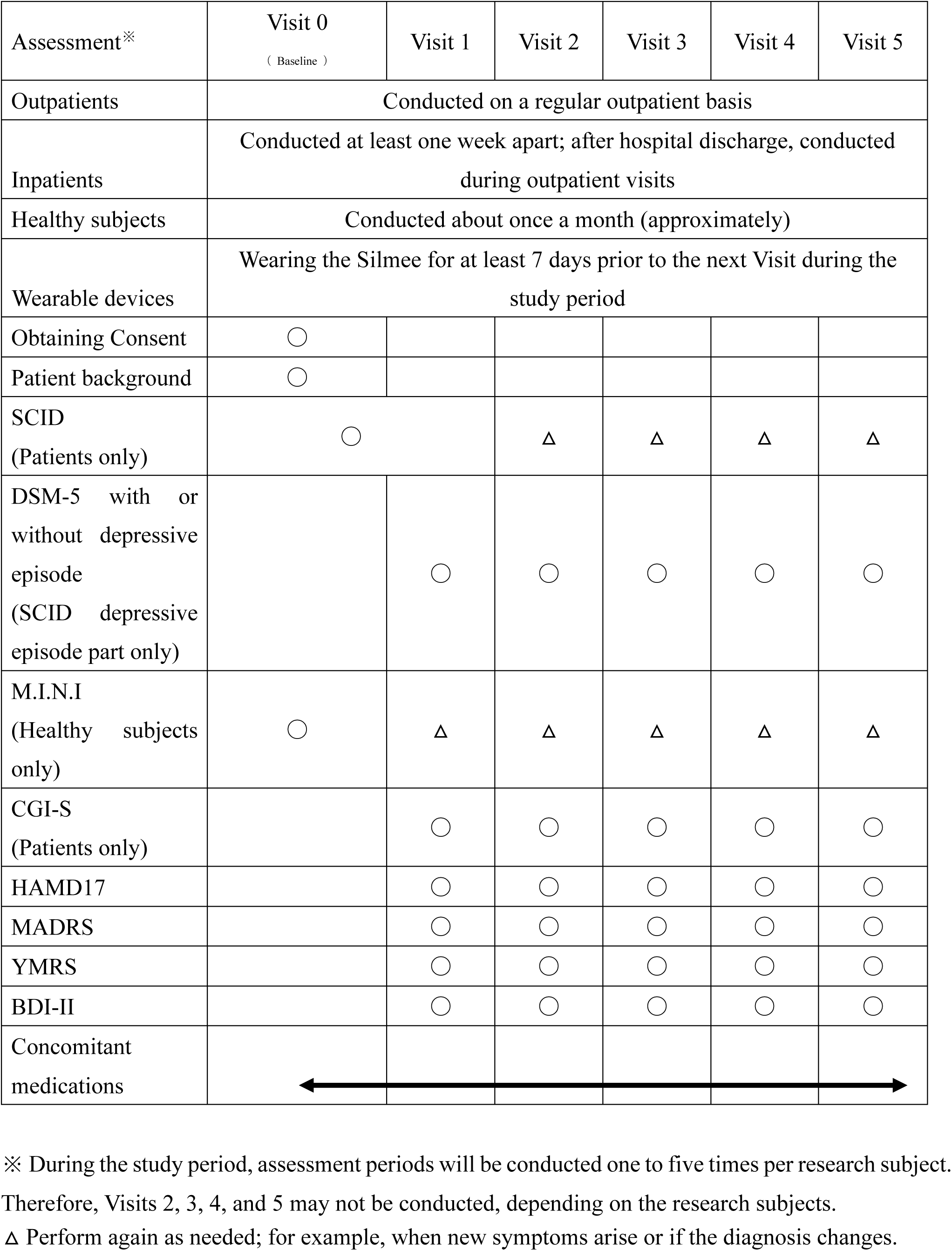
Schedule for data collection and evaluations during the study’s observation period

### 2.4. Primary Outcomes

The primary outcome measures are the machine-learning algorithm’s estimation of whether the patient meets the DSM-5 definition of a depressive episode and the HAMD machine-learning algorithm’s total score estimate.

### 2.5. Secondary Outcomes

Secondary outcome measures include the estimated scores of the HAMD, MADRS, YMRS, and BDI-II sub-items using the machine learning algorithms, and the estimated total score of the combined scores of multiple items. In addition, we will conduct an exploratory analysis using biostatistics and machine learning to examine the relationship between the results of an analysis of structural brain information and brain functional connectivity and the amount of daily life activity and other data obtained from the wearable devices.

### 2.6. Sample Size

In our previous study, we collected 241 datasets over 7 days from 86 patients (45 depressed patients and 41 healthy subjects). In the learning model using XGBoost, the accuracy of identifying symptomatic depressed patients was 76%, and the correlation coefficient with the 17-item version of HAMD for severity assessment was 0.61.^9^ To improve the accuracy of machine learning, as many datasets as possible should ideally be used. However, to minimize the burden on the research subjects and in consideration of feasibility, etc., we created a learning curve based on the accuracy of detection models and severity assessment models using various sample sizes from the previous study mentioned above. As a result, about 500 data sets were estimated to be necessary to achieve a rough target of 90% for the detection accuracy of depressive episodes (Fig. 1). Additionally, about 650 data sets were estimated to be necessary to achieve a severity assessment accuracy corresponding to a correlation coefficient of about 0.85 between the measured and estimated HAMD values (Fig. 2). Furthermore, when the average absolute error from the measured HAMD value was set at 3, about 800 data sets were estimated to be necessary (Fig. 3). These estimates were based on the results of typical and ideal data collections.

**Figure 1.**
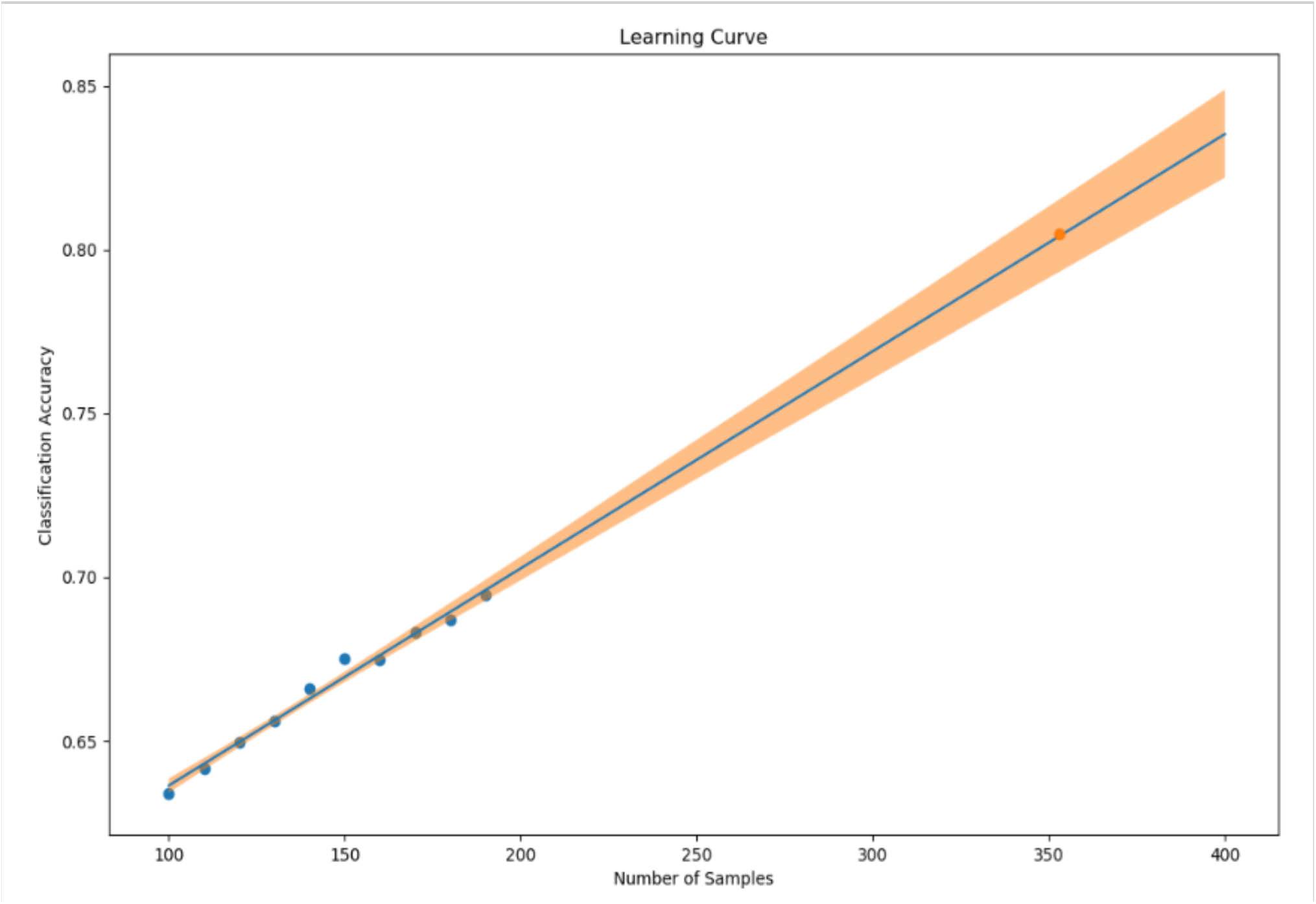
Sample size (X-axis) and mean absolute error between the 17-item version of the HAMD actual and estimated values (Y-axis)

**Figure 2.**
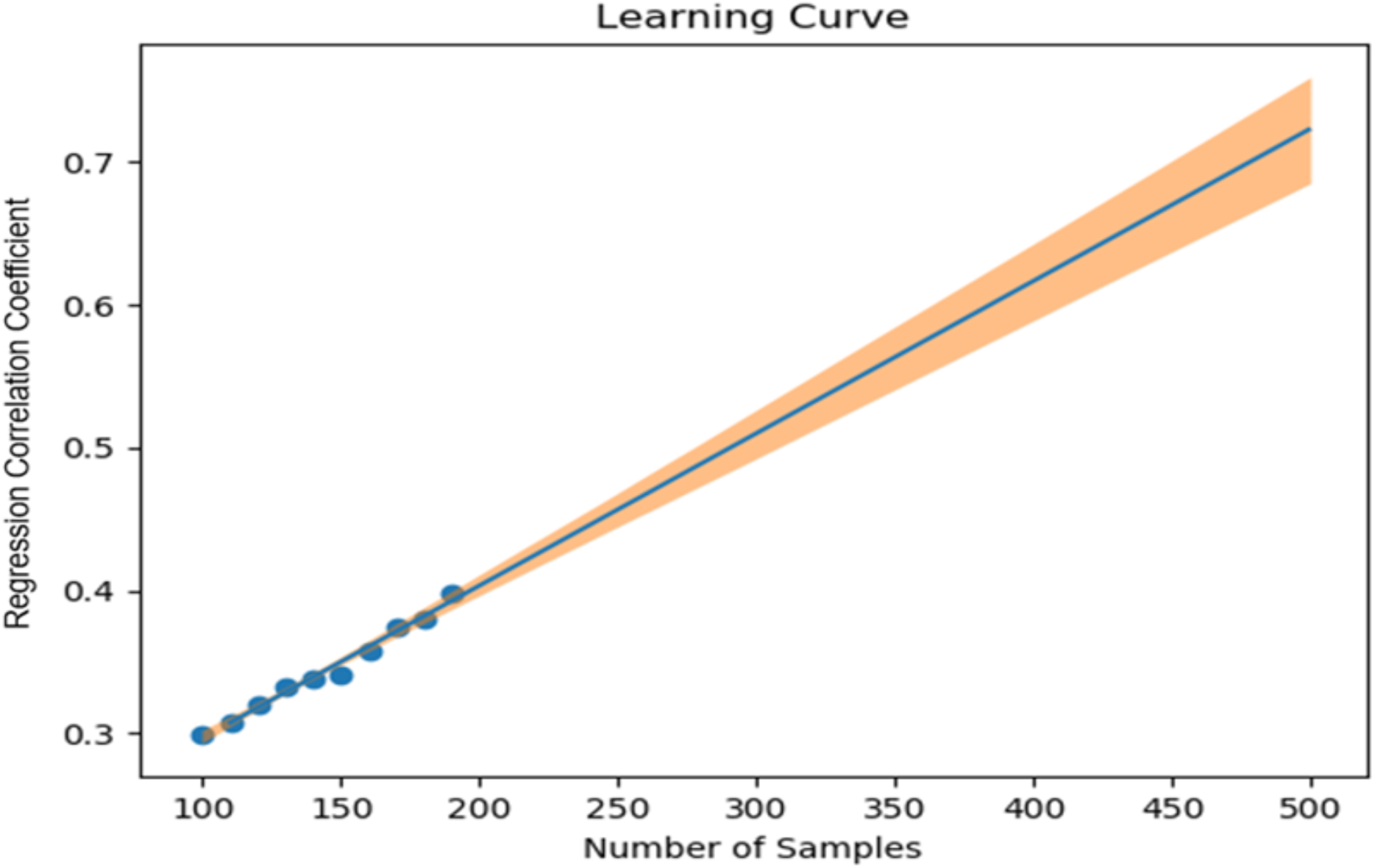
Sample size (X-axis) and correlation coefficient between the 17-item version of the HAMD measured values and the estimated values (Y-axis)

**Figure 3.**
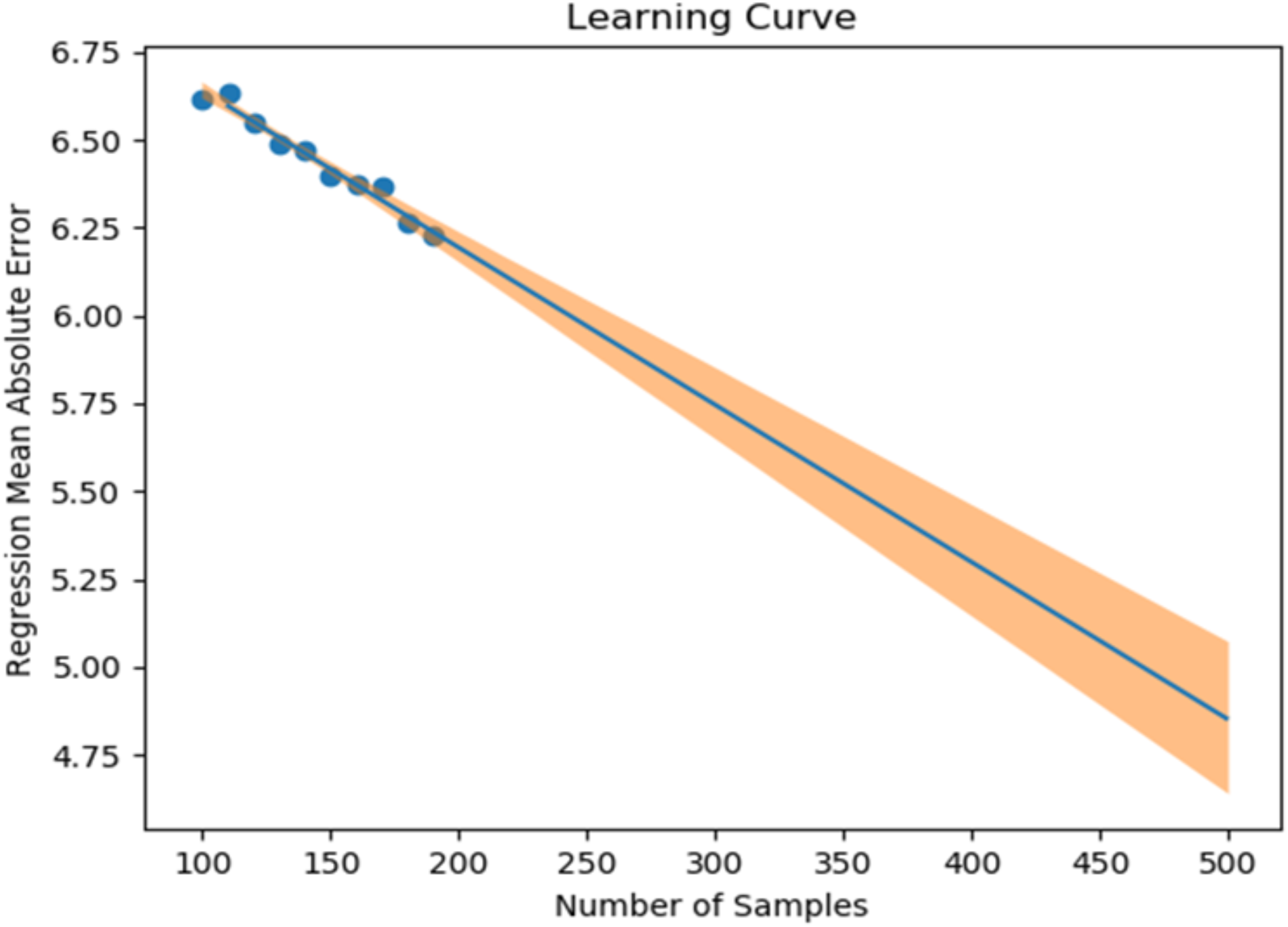
Sample size (X-axis) and mean absolute error between the 17-item version of the HAMD actual and estimated values (Y-axis)

Ideally, a training data set should contain equal numbers of data sets that meet the criteria for depressive episodes and data sets that do not meet the criteria for depressive episodes for detection purposes; for the severity estimation, the data sets should be evenly distributed across a wide range of severities. In practice, however, symptom severity generally declines during follow-up, even if it was severe at baseline, resulting in a large number of data sets with less severe states. For this reason, the severity of the disease cannot be completely equalized. In the proposed study, we plan to collect data from each research subject for up to five measurement periods, if they are willing to participate. To reach the aforementioned target number of data sets, the numbers of patients with each disorder and healthy controls were determined as follows: 220 patients with depression or bipolar disorder, 40 patients with other psychiatric disorders, and 40 healthy subjects, for a total of 300 patients. However, once the target number of cases or half of the data set has been collected, the accuracy of the detection of depressive episodes and the accuracy of the HAMD-17 estimation will be verified, and the target number of cases will be adjusted upward or downward according to an Adaptive Design.

### 2.7. Data Collection and Management

The Silmee W22 (TDK Corporation) wearable device, which is commercially available as a non-medical device, will be used in this study. The Silmee W22 is capable of calculating the pace of walking or running using acceleration data from a 3-axis accelerometer. The data from the Silmee W22 will be transmitted to cloud storage via a dedicated app. The only information that will be uploaded to the cloud will be the device number of the Silmee W22 paired with the dedicated app and the data collected through the Silmee W22. Subjects will be asked to install the dedicated app on their own smartphones and to keep it open or to open it at least once every three days.

In this study, electronic data capture (EDC) will be used to collect information such as the date of consent,patient background, SCID diagnosis name, HAMD, MADRS, YMRS, BDI-II, CGI-S, and M.I.N.I. results, and concomitant medications. We will use “cubeCDMS” from C-Cube Inc. for the EDC.

### 2.8. Statistical Analysis

All research subjects will be included in the analysis. Even if some data for the 7-day measurement period is missing, the data set will be used as training data for machine learning. The data from the Silmee W22 will be denoised and feature-engineered to create a machine learning algorithm that is highly correlated with the presence and severity of depressive episode ratings obtained during researcher-conducted interviews with the subjects. For denoising, we will use techniques such as Continuous Decomposition Analysis (CDA), Discrete Decomposition Analysis (DDA), Gaussian filters, and an adaptive color threshold. These methods are not limited to raw data. If a more suitable method for denoising the raw data becomes available, that method will be used. For feature engineering, we will use autocorrelation, cross-correlation, and other analysis methods to extract temporal structures and patterns for data with time delays. Regarding the feature engineering for heart rate variability, parameters such as SDNN, RMSSD, SDSD, NN50, and Estimated Breath Cycle (EBC) will be used as time-domain features, and low-frequency (LF) and high-frequency (HF) peaks, LF and HF power, normalized LF and HF power, LF/HF ratio, etc. will be used. We will also search for other features that are useful for improving accuracy and will use the optimal methods.

For machine learning algorithms, we will use meta-models such as Support Vector Machine (SVM), Support Vector Regression (SVR), Gradient Boosting Classification/Regression, and so on; alternatively, we will use Deep Learning to find the method that gives the highest accuracy. The significance level will be set at 0.05.

## 3. Results

The trial began in December 2021 and is projected to be completed in June 2026. The trial is registered at jRCT1031210478.

## 4. Discussion

The SWIFT study may lead to the development of medical devices as new diagnostic tools in the field of psychiatry, where biomarkers are scarce. If this technology enables the early diagnosis of depression and can identify signs of recurrence, doctors who do not specialize in psychiatry will be able to refer patients to psychiatrists at an early stage, and occupational physicians will be able to take measures such as reducing workload at an early stage. In addition, the use of this technology by psychiatrists will make it easier to provide measurement-based medical care. As the developed technology will be able to indicate the severity of depression, treatment effects will be easier to detect. Utilizing such technology to clarify symptom changes may lead to improvements in the overall quality of medical care. In addition, patients will be able to monitor their own symptoms. Since previous studies that merely provided information on the status of patients’ symptoms improved patients’ symptoms,^14^ the introduction of medical devices that enable such symptom monitoring may be effective for patient treatment.

The application of AI in medicine has been accelerating year by year, and as of 2020, when this study was planned, 11 AI medical devices have been approved by regulatory authorities in Japan.^12^ However, none of them have targeted psychiatry or have used wearable devices. Similarly, the AI-based medical devices approved by the US FDA as of January 2020 did not include those mounted on wearable devices or those that can diagnose depression.^13^ Thus, the development targeted by the present study is highly novel.

One of the limitations of the learning model to be developed in this study is that it may not be sufficiently accurate to diagnosis or assess the symptoms of severely ill people, since the number of severely ill subjects who are able to participate in this study is expected to be limited because the psychiatric symptoms of severely ill people might prevent them from using wearable devices properly. In addition, elderly people might have difficulty operating wearable devices. Regarding these points, we will strive to incorporate a good balance of ages and illness severities during the recruitment stage.

This study will only recruit Japanese subjects; whether the same diagnostic accuracy can be achieved in other ethnic groups is unclear. In addition, although the information obtained by Silmee is similar to that of wearable devices used in other studies,^15^ the accuracy and content of the information obtained may change in the future as the performances of wearable devices improve. In the future, it would be desirable to mount the system on other wearable devices.

Finally, a certain number of users may have concerns, such as privacy, about the wearable device itself^16^ or may wish to avoid combining wearables with AI.^17^ Taking such concerns and/or preferences of patients into consideration will be needed from an ethical perspective before real-world clinical implementation.

## Data Availability

Not applicable.

## Acknowledgements

The authors are grateful to Ms. Kumiko Hiza and Ms. Hiromi Mikami for supporting the data collection and management in this study.

## Contributions

All authors assisted in the conceptualization of this study. All authors were involved in protocol development. TK was responsible for gaining ethical approval and acquiring funding. TK and SK wrote the first draft of the manuscript. All authors reviewed and edited the manuscript and approved the final version.

## Declaration of competing interests

The author(s) declare the following potential conflicts of interest with respect to the research, authorship, and/or publication of this article:

T. Kishimoto is the president of i2medical LLC and has received consultant fees from Otsuka, Pfizer, and Sumitomo and speaking fee from Banyu, Eli Lilly, Sumitomo, Janssen, Novartis, Otsuka, and Pfizer. T. Horigome is a board member of i2medical LLC and had received consulting fees from FRONTEO Inc. Y. Tazawa is a board member of i2medical LLC. K.C. Liang is a board member of i2medical LLC. N. Koga is a board member of i2medical LLC. G. Miura has received speaking fees from Janssen, Meiji Seika Pharma, Otsuka, and Yoshitomi Yakuhin.

The authors have no other conflicts of interest to declare.

## Ethical approval

The Keio University School of Medicine Ethics Committee approved this study [Approval no. No 20211060, 8 November 2021].

## Funding

This research is supported by the Japan Agency for Medical Research and Development (AMED) under Grant Number 21uk1024003h0001. The funding source did not participate in the design of this study and will not have any hand in the study’s execution, analyses, or submission of results.

Japan Agency for Medical Research and Development (AMED) 20F Yomiuri Shimbun Bldg. 1-7-1 Otemachi, Chiyoda-ku, Tokyo 100-0004 Japan Tel: +81-3-6870-2200, Fax: +81-3-6870-2241, Email: jimu-a.

## Notes

### Clinical Trial

jRCT1031210478

### Author Declarations

The Keio University School of Medicine Ethics Committee gave ethical approval for this work.

